# Recovery from Amblyopia in Adulthood: A Meta-Analysis

**DOI:** 10.1101/2023.05.23.23290409

**Authors:** Madison P. Echavarri-Leet, Hannah H. Resnick, Daniel A. Bowen, Deborah Goss, Mark F. Bear, Eric D. Gaier

## Abstract

**Purpose:** The effectiveness of traditional amblyopia therapies is largely restricted to childhood. However, recovery in adulthood is possible following removal or vision-limiting disease of the fellow eye. Study of this phenomenon is currently limited to isolated case reports and a few case series, with reported incidence ranging from 19-77% ^1–5^. We set out to accomplish two distinct goals: (1) define the incidence of clinically meaningful recovery and (2) elucidate the clinical features associated with greater amblyopic eye gains.

**Methods:** A systematic review of 3 literature databases yielded 23 reports containing 109 cases of patients ≥18 years old with unilateral amblyopia and vision-limiting fellow eye pathology.

**Results:** Study 1 revealed 25/42 (59.5%) of adult patients gained ≥2 logMAR lines in the amblyopia eye after FE vision loss. The overall degree of improvement is clinically meaningful (median 2.6 logMAR lines). Study 2 showed that for cases with amblyopic eye visual acuity improvement, recovery occurs within 12 months of initial loss of fellow eye vision. Regression analysis revealed that younger age, worse baseline acuity in the amblyopic eye, and worse vision in the fellow eye independently conferred greater gains in amblyopic eye visual acuity. Recovery occurs across amblyopia types and fellow eye pathologies, although disease entities affecting fellow eye retinal ganglion cells demonstrate shorter latencies to recovery.

**Conclusions:** Amblyopia recovery after fellow eye injury demonstrates that the adult brain harbors the neuroplastic capacity for clinically meaningful recovery, which could potentially be harnessed by novel approaches to treat adults with amblyopia.

## Introduction

Amblyopia is a prevalent visual disorder resulting from abnormal visual experience during early development^6–8^. The long-standing and current treatment standard involves occlusion via patching or pharmacologic cycloplegia of the fellow eye (FE). While most children with amblyopia experience visual acuity (VA) gains in the amblyopic eye (AE) following standard therapy, improvement is often incomplete and restricted to cases where therapy is initiated before age 8^9, 10^. As such, treatment of amblyopia is widely considered futile in adults in routine ophthalmic practice.

There is extensive evidence that at least partial recovery from amblyopia can occur in experimental animal models late in the critical period (when simple occlusion-type interventions fail) and beyond^11–14^. For instance, experiments in mice and cats show that temporary inactivation of retinal ganglion cell (RGC) activity in both eyes or just the FE establishes conditions that enable the rapid and durable recovery from amblyopia^15–17^. This intervention is effective at ages when reversed occlusion (analogous to patch therapy in humans) fails to promote sustained recovery^16, 17^. Enucleation and retinal lesioning similarly promote recovery in visual cortical responses to stimulation of AEs of adult cats, monkeys, and mice^18–22^. Direct limitation of RGC activity is a common theme distinguishing the interventions that successfully drive recovery in older animals from those that do not. Thus, the pre-clinical, experimental basis for amblyopia recovery in adulthood is strong and broad, but comprehensive supporting evidence in humans is needed.

Analogous observations of recovery from amblyopia triggered in human patients who suffer vision-limiting FE disease or loss spans decades of clinical literature. To date, there have been 5 dedicated studies examining amblyopia recovery following FE vision loss; 3 focus on specific FE disorders (ischemic optic neuropathy^1^, age-related macular degeneration^2^, uveal melanoma^3^), and 2 include all causes of FE visual loss^4, 5^. Additionally, numerous isolated reports describe the clinical courses of 1-3 patients in various clinicopathologic contexts. Despite this substantial literature, key features of this important phenomenon, which may hold the key to reversal of amblyopia in adulthood, remain undefined.

To begin to fill this knowledge gap, we integrated and analyzed all available clinical descriptions of AE VA recovery in the context of FE disease in adults. We aimed to (1) describe the prevalence and magnitude of recovery and (2) answer whether specific clinical metrics influence the magnitude of AE VA gains. We tested the hypotheses that direct FE RGC injury and absence of strabismus would confer greater AE VA improvement.

## Methods

### Database Search

A systematic search of PubMed (1946-), Embase (1947-), and Web of Sciences Core Collection (1900-) databases was conducted using combinations of the following key words: “amblyopia”, “recov-”, “revers-”, “loss”, “injury”, “visual impairment”, “fellow”, “fixating”, “dominant”, “good”, “contralateral”, “eye”. Initial searches were designed and executed by HHR and DG. Final searches were run on December 21^st^, 2021. A total of 1660 unique abstracts (deduplication from 2925 using Endnote X7.8) were returned. Covidence software (https://www.covidence.org) was used to manage the review process. Each unique abstract was screened by two independent team members, and conflicts were resolved by EDG. PRISMA guidelines were used for documentation^23^.

### Article inclusion

Abstracts were screened based on the following inclusion criteria: cases of human adults (≥18 years of age) with amblyopia and FE vision loss; provides individual patient-level data; provides at least 2 AE VA measurements at different timepoints.

### Article exclusion

Articles focused on basic neuroscience/experimental techniques/interventions were excluded. Reports that were not published, were not primary articles (e.g., book chapters, lectures, abstracts, presentations, reviews) or were not available in English were excluded.

### Literature analysis

After abstract screening, 35 articles were reviewed in detail, and 17 ultimately met all inclusion and exclusion criteria. Bibliographies of each included report were reviewed for additional potentially relevant reports not captured in our database searches. All referenced studies were collected and reviewed in detail, yielding 6 additional reports meeting criteria. In total, 23 reports (comprised of 109 unique cases) were included for analysis (Supplemental Figure 1).

### Data Extraction

Each report was comprehensively and systematically examined. We extracted all parameters of interest for further analysis for each patient when available, including:

- Initial AE VA before the injury, if available, or else at the time of first evaluation after FE vision loss.
- Best measured AE VA after FE vision loss.
- Time interval from FE visual loss to best AE VA measurement (in months). If the time fell in the middle of the month or the exact length of time could not be determined, the time was rounded to the next month. If months were not provided for each timepoint, the difference in years was multiplied by 12. Times less than 1 month were rounded to one month.
- FE VA nadir, defined as the worst reported FE VA after onset of FE visual loss
- Patient age at the time of FE loss; age was binned as 18-40, 41-60, and ≥61 years for analysis.

All VA measures were converted to the logarithm of the minimum angle of resolution (logMAR) scale and rounded to the nearest 0.02 unit (1 letter). AE VA logMAR lines of improvement was calculated from best-initial AE VA logMAR values.

### Analysis

We conducted 2 separate goal-directed studies. Study 1 aimed to determine the incidence and magnitude of AE VA improvement. To minimize the influence of reporting bias, only series reporting consecutive subjects with patient-level data within a defined study period (agnostic to whether recovery occurred for each individual patient) were included (42 cases, 3 series). Study 2 aimed to describe the clinical features associated with AE VA gains and thus focused on only cases with measurable (>0.0 logMAR) improvement (101 cases, 23 reports). Among these, 55 cases specified FE VA measurements (to assess the influence of FE visual loss magnitude), 37 cases specifically reported on the presence/absence of strabismus (for segregation by amblyopia type), 38 cases had a clearly documented cause of FE vision loss (for segregation by direct RGC involvement), and 80 cases specified the timeline from FE vision loss to key AE VA measurement in follow-up (to assess temporal rate of recovery) (Figure 1).

**Figure 1:**
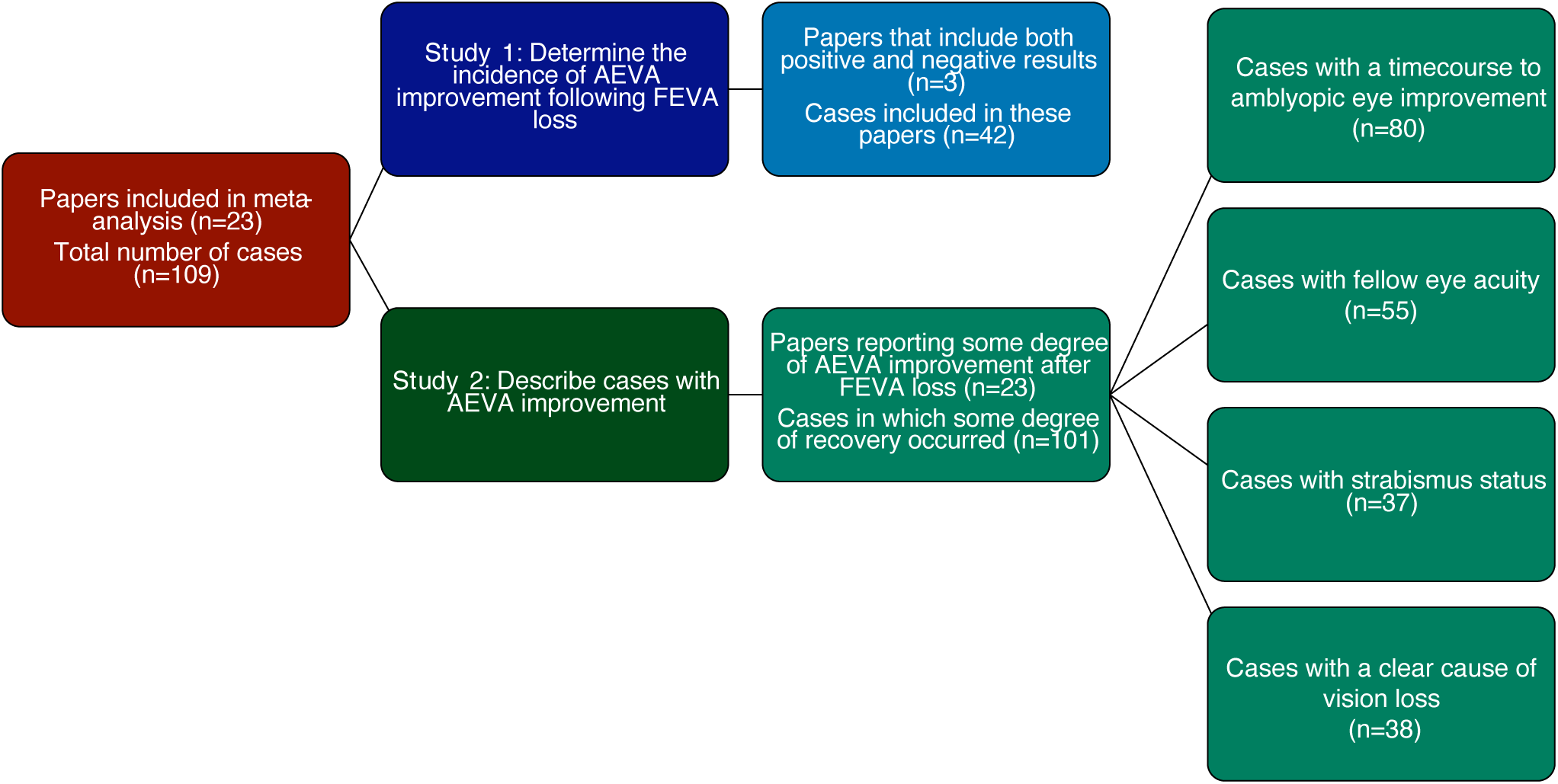
Inclusion criteria and case selection for Studies 1 and 2. 109 cases from 23 papers met our initial inclusion criteria. To determine the incidence of AE VA improvement following FE VA loss for Study 1, we only included papers that reported both positive and negative results to minimize the influence of reporting bias. Only 3 of the 23 papers reported negative and positive results; the remaining 20 papers only reported positive cases. Thus, only the 42 cases from these 3 papers were included in Study 1. In Study 2, we focused only on cases where some degree of AE VA improvement was observed. 101 cases from 23 papers met this criterion. Some analyses within Study 2 were restricted to cases in which specific data were available: time course to maximal AE (n=80), FE VA nadir (n=55), strabismus status (n=37), and etiology of FE vision loss (n=38).

### Statistics

Normality of data was assessed with the Kolmogorov-Smirnov test. Direct comparisons between two normally distributed groups employed a two-tailed t-test, whereas those involving one or more groups that were not normally distributed employed a Mann-Whitney test. In the case of comparing initial to best AE VA values (which were not normally distributed) a Wilcoxon matched-pairs test was used to assess whether changes in individual case AE VA values changed significantly. A univariate multiple regression was performed (SPSS, IBM, Armonk, NY) with input variables: age, baseline AE VA, FE VA nadir. The output variable was AE VA (logMAR lines) of improvement. A modified Breusch-Pagan test determined that the data were heteroskedastic (*p*=0.006). To control for this, robust standard errors calculation (HC3) was used in calculations of *p*-values in the linear regression. Bias in the linear model based on whether data came from a paper with a small sample size (≤3 cases) or a larger sample size (>3 cases) was assessed by comparing the effect size of each of the independent variables^24, 25^. The threshold for statistical significance was set at *p*<0.05 in all cases.

## Results

### STUDY 1: Incidence of clinically meaningful recovery

Of the 109 reports revealed by our review, 23 were isolated case reports or small (≤3 cases) series. To mitigate the effects of reporting bias through underrepresentation of cases without recovery, we analyzed the 42 cases originating from 3 reports containing >3 cases that were agnostic to the occurrence of recovery and included patient-level data.

Among these 42 cases, the median change in AE VA was 2.60 logMAR lines [IQR:0.95-4.00]. Eight (19.0%) had no AE VA improvement. Among the remaining cases, 34 (81.0%) had measurable AE VA improvement, 32 (76.2%) gained ≥1 logMAR line, 25 (59.5%) gained ≥2 logMAR lines, and 21 (50.0%) cases gained ≥3 lines (Figure 2). These findings demonstrate a high incidence of clinically meaningful recovery among adults after loss of vision in the FE.

**Figure 2:**
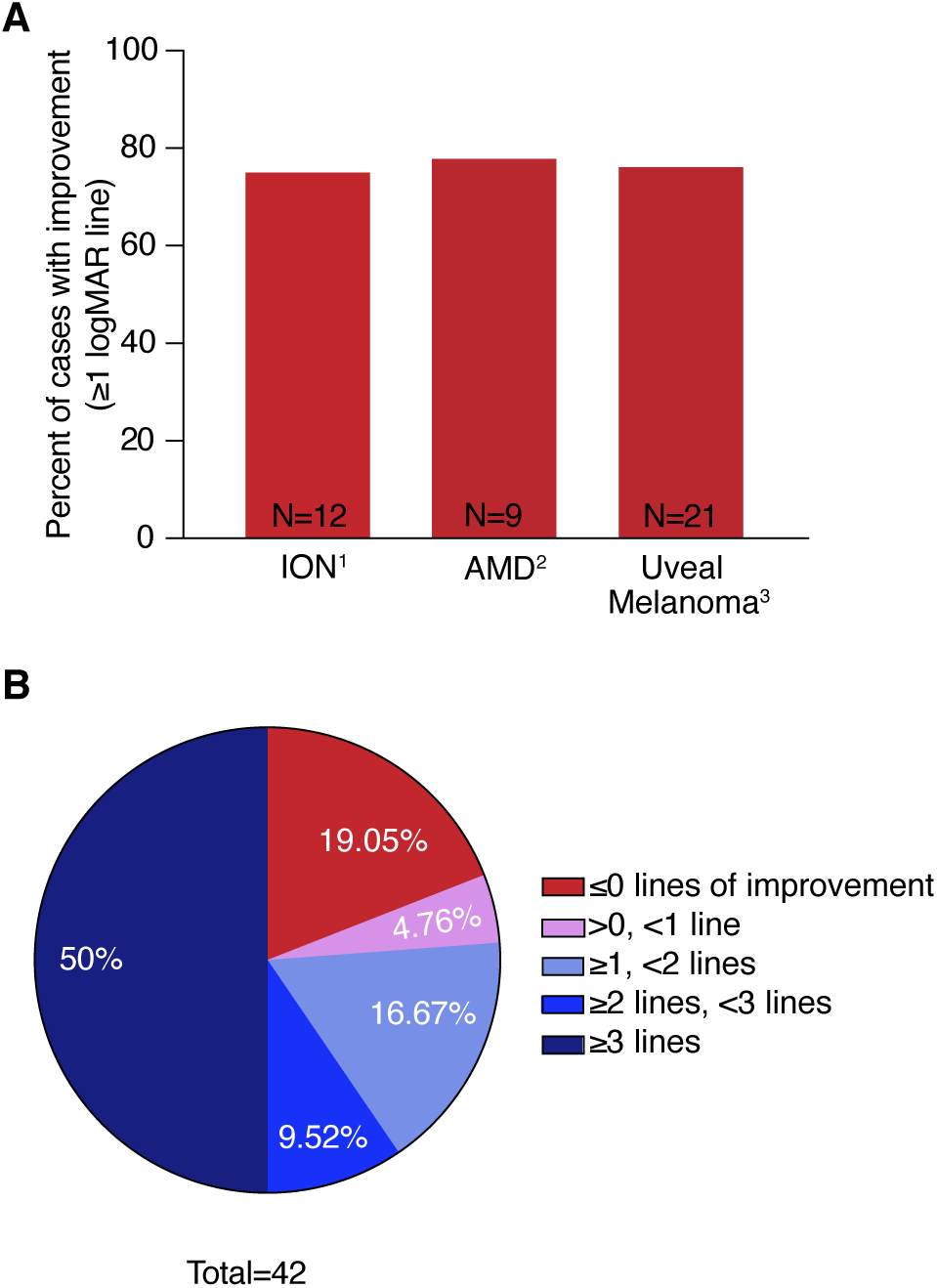
Following fellow eye vison loss, the majority of adults improve visual acuity in the amblyopic eye. **(A)** Three dedicated studies met our inclusion criteria for the purposes of Study 1, with a similar incidence of recovery observed in each. **(B)** Pie chart illustrating how many cases had various degrees of AE VA improvement.

### STUDY 2: Features influencing amblyopic eye recovery

To identify clinical factors associated with greater AE VA improvement after FE vision loss, we narrowed the 109 cases in our review to the 101 who demonstrated AE VA gains. Among these, median initial AE VA was 1.00 logMAR [IQR: 0.50-1.00)]. The median best AE VA following FE vision loss was 0.22 logMAR [IQR: 0.10-0.42] (Figure 3A, B). The median change in AE VA following loss of vision in the FE was 5.0 logMAR lines of improvement, which was significant via a Wilcoxon matched-pairs test (*p*<0.0001). 84/101 (83.2%) of patients exhibited ≥3.0 logMAR lines of improvement (Figure 3C). Median time from FE vision loss to maximum AE VA was 12.0 months [IQR: 6.25-24.00] (Figure 3D), though this likely reflects a frequent timepoint for scheduled follow up.

**Figure 3:**
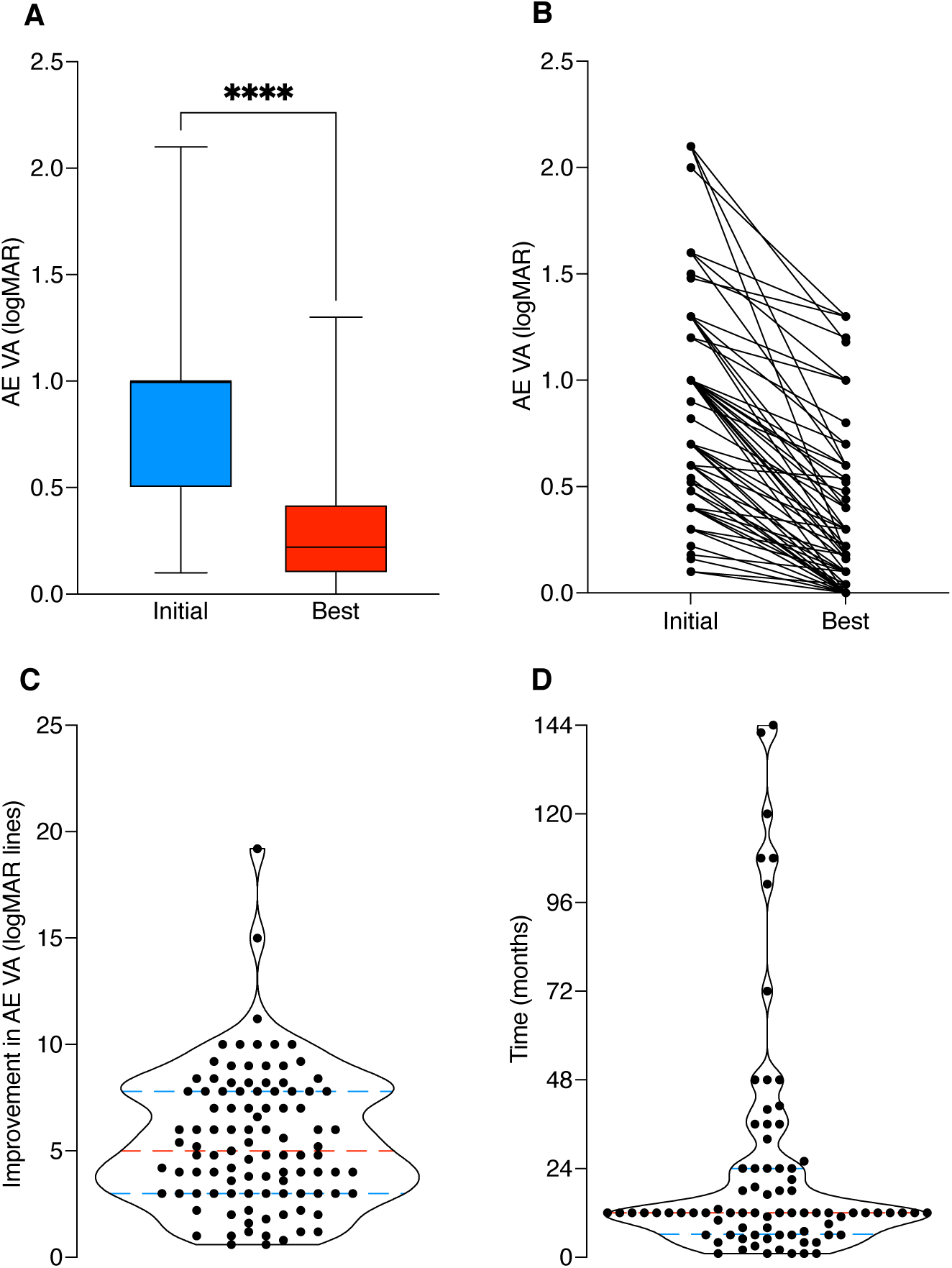
For most adult amblyopes who improve amblyopic eye visual acuity following loss of vision in the fellow eye, recovery occurs within 12 months. **(A)** Box plots depicting median logMAR VA (black line) at baseline (blue) and maximal AE VA following FE vision loss (red) (**** *p*<0.0001). The top and bottom borders of the box indicate quartiles, and whiskers indicate minimum and maximum values (n=101). **(B)** Individual case data of baseline and maximal AE VA (n=101). **(C)** Violin plot depicting the distribution of AE VA improvement for all cases (n=101). The red line depicts the median, and blue lines depict quartiles. **(D)** Violin plot depicting the distribution of time from FE vision loss to maximal AE VA (n=80). The red line depicts the median, and blue lines depict quartiles.

### Linear modeling of clinical feature influence on recovery

A linear model was constructed to assess the relationship between binned age, initial AE VA, and FE VA nadir with logMAR lines of improvement for the AE following FE vision loss. This analysis was restricted to the 55 available cases with recorded FE VA data. Younger age was associated with greater AE VA gains with an effect size of 1.7 logMAR lines for each younger age bin (*p*=0.002). More severe amblyopia was associated with greater AE VA gains with an effect size of 4.9 logMAR lines of improvement (*p*<0.001). Worse FE VA nadir correlated with greater AE VA gains, with an effect size of 1.0 logMAR lines of improvement (*p*=0.005) (Figure 4).

**Figure 4:**
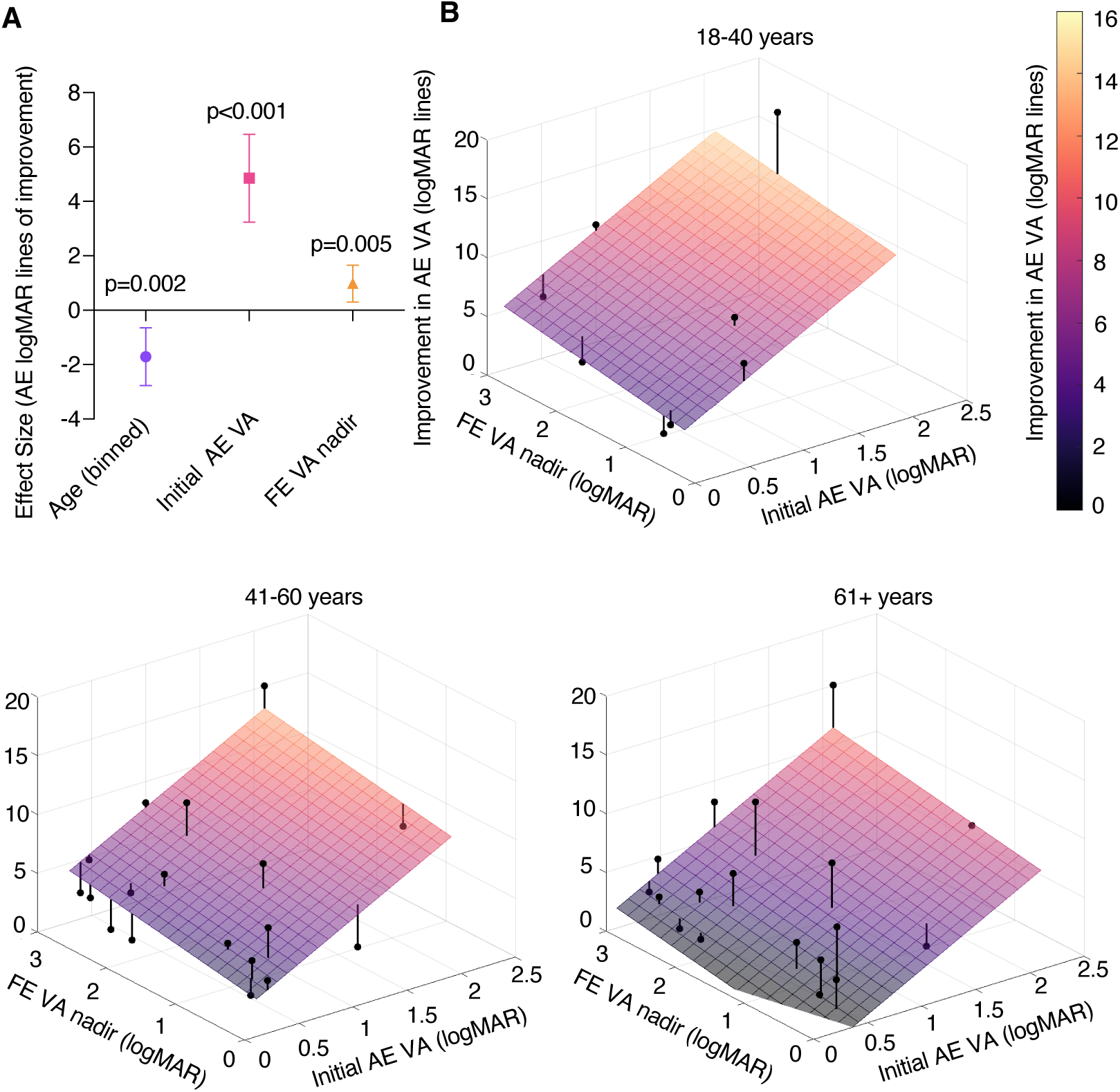
Linear modeling reveals younger age, worse initial amblyopic eye visual acuity, and worse fellow eye visual acuity following vision loss are independently associated with greater amblyopic eye visual acuity improvement. **(A)** Effect sizes of significant factors identified by multiple linear regression using robust standard errors. Bars represent 95% confidence intervals. **(B)** Mesh grids of predicted values and calculated slope from the linear model segregated by age bins: 18-40 (left), 41-60 (center), 61+ (right). Individual case AE VA improvement values are represented by black dots and lines connect to the plane of best fit.

### Hypothesis-driven categorical clinical feature influence on recovery

Amblyopia etiology was unavailable for 64 patients, so they were not included in this analysis. 18 patients had a history or presence of strabismus, and 19 patients did not. There was no significant difference in AE VA improvement between those with (median: 4.00 [IQR: 1.95-8.40]) versus those without (median: 3.90 [IQR: 2.20-6.00]) strabismus (*p*>0.9; Mann-Whitney test). (Figure 5A).

**Figure 5:**
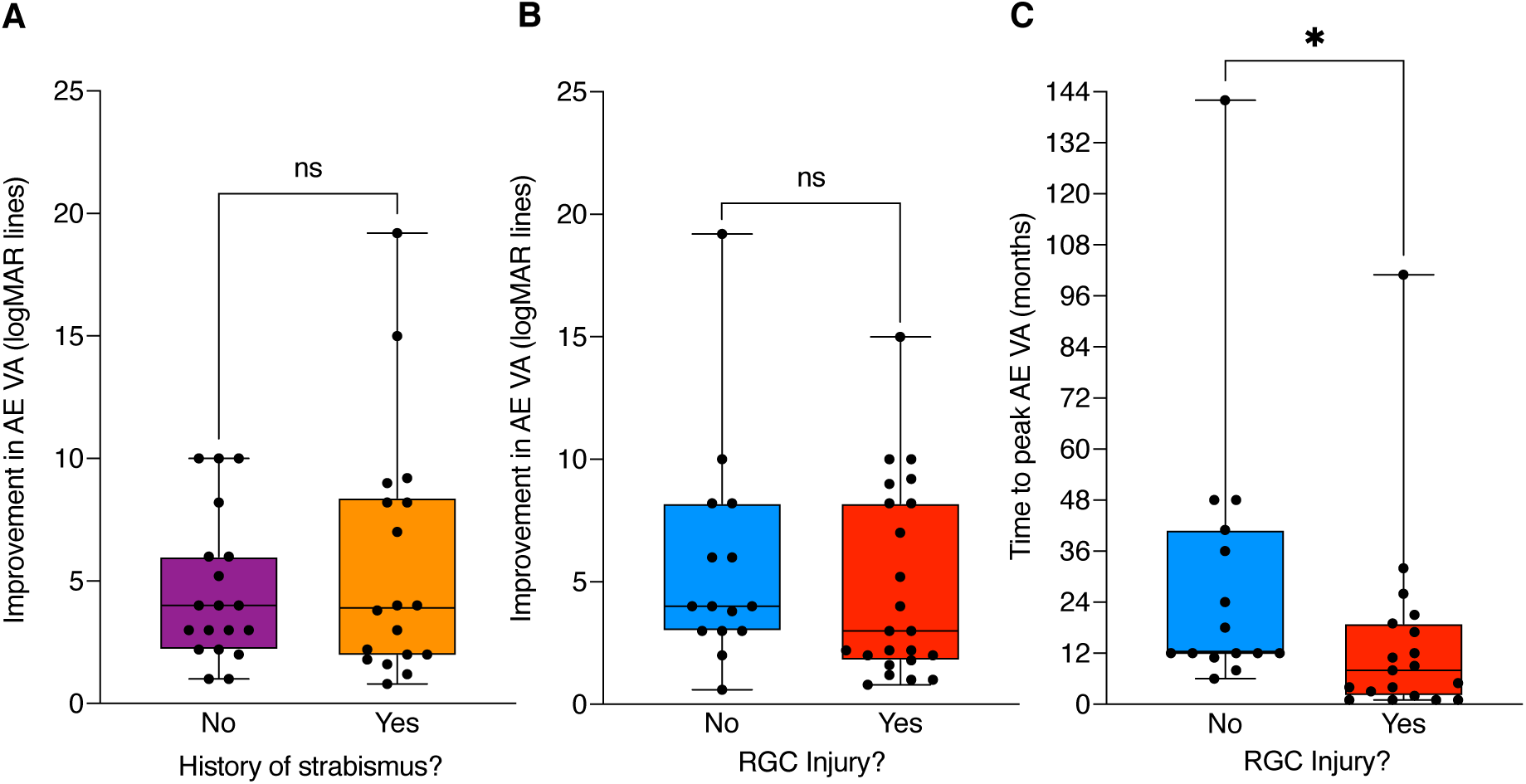
Testing the influence of categorical clinical features of amblyopia or fellow eye vision loss on the degree of amblyopia eye visual acuity recovery. **(A)** Box plots depicting median logMAR line VA improvement (black line) according to strabismus status (ns *p*>0.9; Mann-Whitney test). Top and bottom borders of the boxes indicate quartiles and whiskers indicate range. **(B)** Box plots depicting median logMAR line VA improvement (black line) according to FE pathology involvement of retinal ganglion cells (RGCs) (ns *p*>0.3; Mann-Whitney test). **(C)** Box plots depicting median time to maximal AE VA (black line) according to FE pathology involvement of RGC (* *p*=0.0124; Man-Whitney test).

Of the 101 cases found through our systematic literature search, 38 had clear documentation relating to the cause of FE vision loss where direct involvement of RGCs could be ascertained. Of those 38 cases, 22 cases of FE vision loss with RGC involvement and 16 cases without RGC involvement. There was no significant difference in AE VA improvement between cases with (median: 3.00 [IQR: 1.80-8.20)]) and without (median: 4.00 [IQR: 3.00-8.20]) RGC injury (*p*>0.3, Mann-Whitney test). Comparing rate of recovery, those with RGC injury saw faster improvement in AE VA (median: 8 months [IQR: 2-19]) than those without (median: 12 months [IQR: 12-41]; *p*=0.0124, Mann-Whitney test) (Figure 5B, C)

### Post-hoc assessment of study bias

Our systematic review returned reports that contained larger case series with >3 cases (n=5) and case reports/smaller case series (≤3 cases) (n=18). Because larger case series contributed 78/101 (77.2%) cases, we asked whether this distribution could influence the results. There was no significant difference between AE VA gains between those contributed by larger case series (median: 4.80 [IQR: 3.00-7.00]) and small case series/reports (median: 7.00 [IQR: 3.90-8.60]; *p*=0.098, Mann-Whitney test) (Supplemental Figure 2A). Confidence intervals for each case series and the composite of small series and isolated reports overlap the median of 5.0 logMAR lines of AE VA improvement for the overall sample without systematic bias trends (Supplemental Figure 2B). We assessed whether the linear clinical factors found to influence AE VA improvement differed systematically across studies. There were no significant differences in regression weights between groups (*binned age* Z=0.056, *p*>0.9; *initial AE VA* Z=1.465, *p*>0.14=; *FE VA nadir* Z=0.704, *p*>0.4), suggesting neither report type imparts measurable bias on regression weights for linear clinical factors. Thus, there was no clear evidence of bias imparted by report type or size on the primary outcome measure in Study 2.

## Discussion

Although conventional occlusion therapy is not effective in adults with amblyopia, gains in AE VA following FE vision loss is common. Integration and analysis of reported cases demonstrate that the adult visual system retains the capacity for plasticity necessary for recovery from amblyopia. These results show that 60% of adults with amblyopia who experience FE vision loss will gain at least 2 lines of AE VA. Amblyopia treatment trials in children <8 years of age reporting an average of 2-3 logMAR lines of improvement following 3-6 months of occlusion treatment suggest that the prevalence of recovery is substantial^8^. By comparison, only 25% of 13–17-year-olds (and 47% of treatment naïve 13–17-year-olds) gain ≥2 lines of AE VA following 2-6 hours of daily patching^26^. While randomized control trials in adults are not available, this comparison suggests that the rate of AE VA improvement seen after FE vision loss markedly exceeds conventional occlusion therapy.

We focused our study of clinical factors on cases with some degree of AE VA improvement to avoid over-representation of recovered cases by inclusion of isolated reports. In this sample, we found that improvement is generally observed within 12 months following FE vision loss, and further gains after this time point are less common. Most amblyopia is associated with anisometropia, strabismus, or both^27^. A history of or presence of strabismus did not influence the degree of AE VA recovery in our study despite more blunted gains^28^ and higher rates of regression^29^ in conventionally treated strabismic compared to anisometropic amblyopia. Given that previous research indicates that temporary inactivation of FE RGCs promotes recovery from amblyopic rearing in mice and cats, we postulated that direct RGC involvement may be required to enable the plasticity necessary to drive recovery in older age, but our results suggest this is not necessarily the case^15, 16^. We noted comparable rates of recovery in outer retinal disorders such as age-related macular degeneration^2^ and uveal melanoma^3^ as with ischemic optic neuropathy^1^. FE injury directly involving RGC damage did not affect the magnitude of AE VA gains but did confer a faster rate of recovery. Though the current analyses focus on injuries involving damage to RGCs, future work could potentially compare pre-retinal (e.g. corneal, lenticular) and retinal/optic nerve disorders.

It is well-documented that the success of amblyopia treatment depends on patient age, and that adults generally do not respond to conventional amblyopia treatment methods^8, 10, 11, 30, 31^. Although our analysis was restricted to adult patients, we were curious whether patient age at the time of FE loss would be a factor associated with degree of AE VA improvements. Linear regression analysis showed that younger age, greater severity of vision loss in the FE, and worse initial AE VA independently influenced the degree of improvement in the AE VA. These results may be applied in clinical contexts to prognosticate the degree of AE recovery in new cases of FE vision loss.

Our study has several limitations, mainly stemming from factors inherent to the nature of meta-analyses. The availability and reliability of data could not be controlled in this systematic literature review spanning many decades, journals and report types. Subgroup analyses and linear models engaged fractions of our sample because of inconsistencies in reported details between cases and reports, and thus there is some susceptibility to bias within our sample. This underscores the importance of our post-hoc analysis, which demonstrated no systematic biases of report type on either the primary outcome measure or the factors found to be influential through linear modeling. Despite these inherent limitations, the relative infrequency of this clinical problem (owing to a need for two separate ophthalmic conditions in each eye: amblyopia and vision-limiting FE pathology) necessitates meta-analysis as the only feasible means to test our hypotheses. However, advancement of means to build and integrate large medical databases could provide another means to study this phenomenon in the future, provided that the minimal patient-level details such as laterality are available.

The Bienenstock, Cooper, and Munro (BCM) theory of metaplasticity^32^ provides a useful conceptual framework to understand how FE vision loss can drive AE recovery^14^. The BCM theory posits that the threshold for cortical synaptic potentiation changes as a function of the history of integrated post-synaptic neuronal activity. In this clinical context, the reduction in visual cortical activity following FE vision loss lowers the plasticity threshold to enable potentiation of synapses serving the AE, ultimately improving AE visual function. Of course, permanent FE vision loss is not a viable treatment approach. However, according to this theoretical model, even temporary silencing of the FE may be sufficient to enable synapses serving the AE to grow in strength and, importantly, maintain these improvements even when the FE activity returns to normal. Understanding the molecular, cellular and circuitry contributions to metaplasticity^14, 33–35^ may lead to novel amblyopia therapies in adults. Experimental elucidation of the relevant mechanisms through animal models will serve to refine potential therapeutic targets whereby the plasticity that enables recovery from amblyopia in adulthood could one day be harnessed.

## Data Availability

All data produced in the present work are contained in the manuscript

## Acknowledgements

Hui Li, PhD candidate in biostatistics at the Harvard T.H. Chan of Public Health, for statistical consultation.

---

**Supplemental Figure 1: Method for collection and inclusion of relevant reports and cases.**

A systematic search of PubMed (1946-), Embase (1947-), and Web of Sciences Core Collection (1900-) databases was conducted and returned 2925 papers. After deduplication, 1660 abstracts were screened by two members of the team for relevance. 35 papers were sought for retrieval and the relevance was assessed based on the article in its entirety. We ultimately determined that 17 of these papers met our eligibility criteria for inclusion in the meta-analysis. Eligible papers then had their individual bibliographies reviewed. Non-duplicate references were retrieved, and 39 additional sources were screened for relevance. 10 ultimately were reviewed in full, and 6 of these met eligibility criteria and were included in the final meta-analysis. In total 23 reports comprised of 109 cases were included. PRISMA guidelines were used for this documentation ^23^.

**Supplemental Figure 2: Assessing potential bias in primary outcome measure (logMAR lines of recovery) by report type. (A)** AE VA (logMAR lines) improvement segregated by report type (ns *p*=0.098; Mann-Whitney test). Error bars indicate 95% confidence interval. Black dotted line indicates the median (y = 5.0 logMAR lines of AE VA improvement) for all cases, regardless of report type. **(B)** Forest plot comparing the median logMAR lines of AE VA improvement (black dots) with 95% confidence intervals (error bars) for all case series containing >3 cases individually and pooled case reports and case series containing ≤3 cases. The number of reported cases and authors for each report of case series comprise the y-axis labels. Red dotted line (x = 5.0) indicates the median logMAR lines of AE VA improvement for all cases, regardless of report type.

**Supplemental Table 1: Summary of individual case details**

A table of all included cases and factors analyzed. Author/Paper: yellow indicates this case was included in Study 1, and grey indicates it was not. Lines of improvement: green indicates this case was included in Study 2, and red indicates it was not. Time to best (months) and Worst FE VA (logMAR): orange indicates this information was available and included in the analysis, and grey indicates it was not. Binned age: pink indicates patients aged 18-40 (group 1), yellow indicates patients aged 41-60 (group 2), and green indicates patients aged 61+ (group 3). History of strabismus: orange indicates patients with a history or presence of strabismus, purple indicates patients without a history or presence of strabismus, and grey indicates patients that were not included in this analysis (either because this information was not available or because the patient did not show AE VA improvement). RGC Damage: red indicates patients with RGC damage, blue indicates patients without RGC damage, and grey indicates patients excluded from this analysis because the RGC status was unclear.

