## Supplementary figures and images for "Recovery from Amblyopia in Adulthood: A Meta-Analysis"

### Supplemental Figure 1

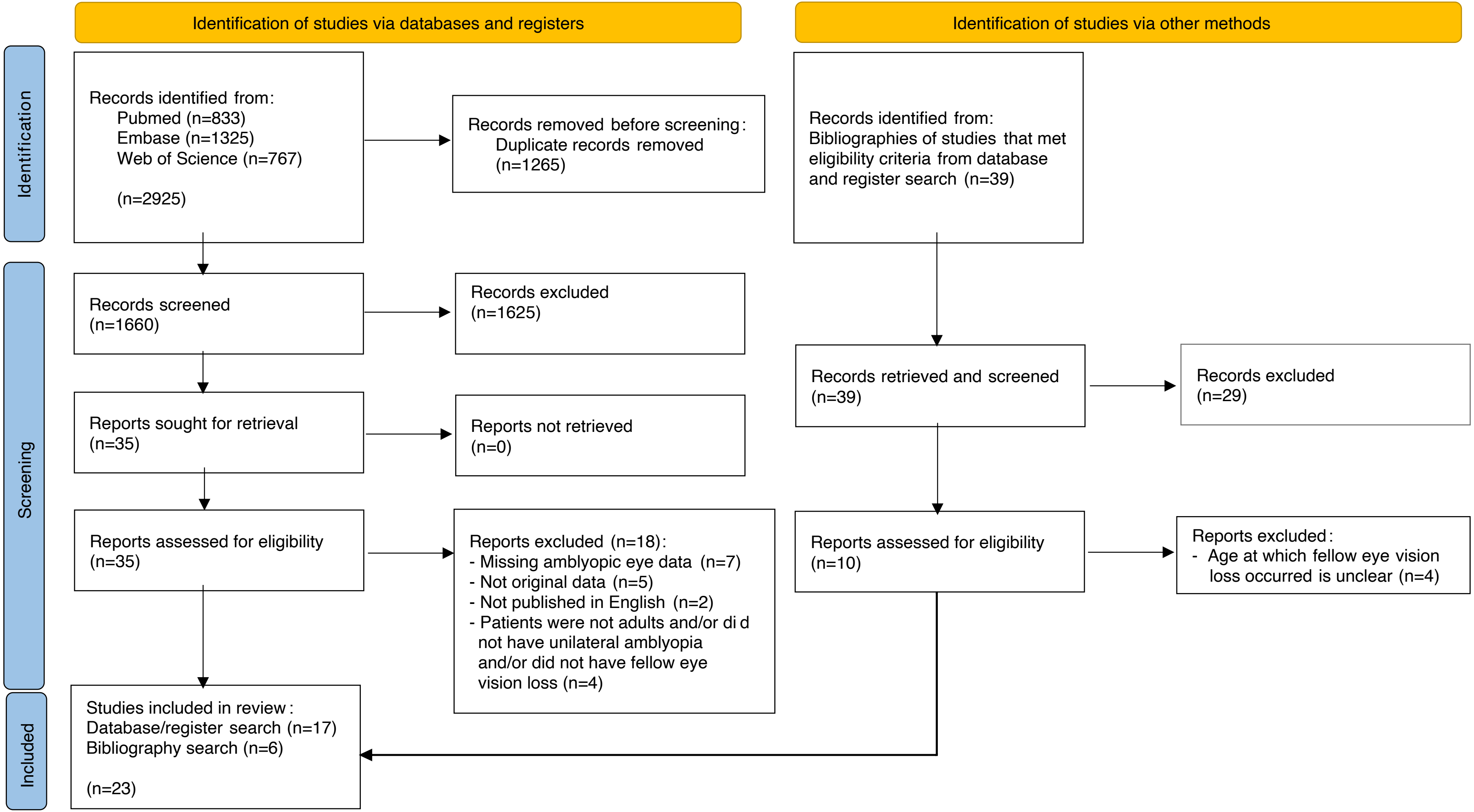

### Supplemental Figure 2

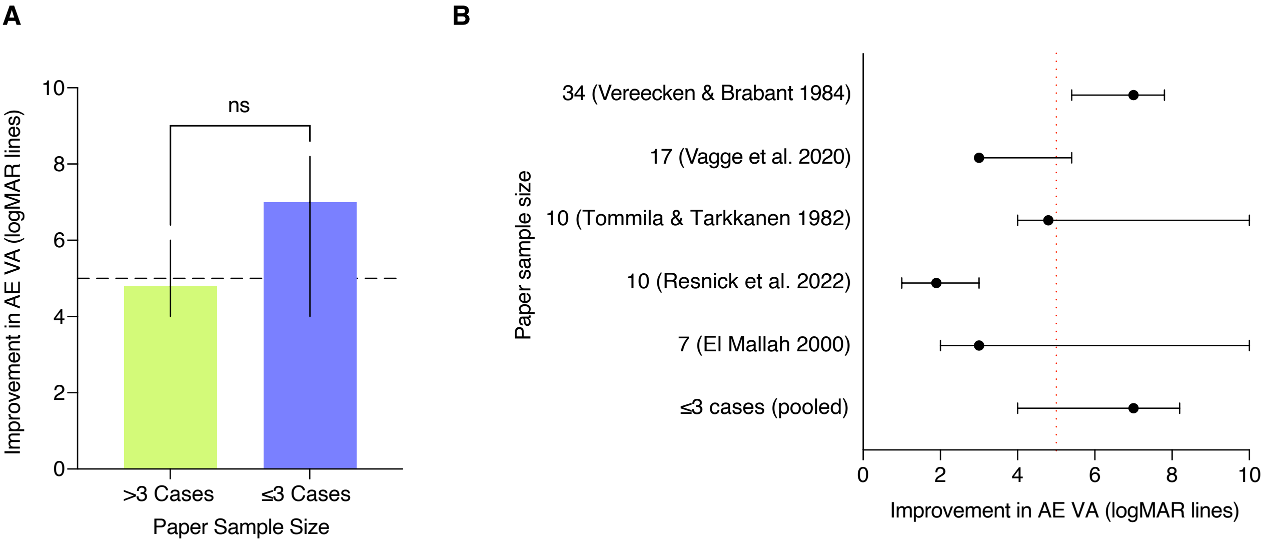
