## Supplemental Table 1 for "Recovery from Amblyopia in Adulthood: A Meta-Analysis"

| Subject # | Author/Paper | Initial AE VA (logMAR) | Best AE VA (logMAR) | Lines of improvement | Time to best (months) | Worst FE VA (logMAR) | Age at FE vision loss | Binned Age | History of strabismus? | RGC Damage |
|---|---|---|---|---|---|---|---|---|---|---|
| 1 | Resnick, H. H., et al. (2022). "Partial Recovery of Amblyopia After Fellow Eye Ischemic Optic Neuropathy." Journal of Neuro-Ophthalmology: 10.1097. | 1.48 | 1.30 | 1.80 | 4 | 0.00 | 76 | 3 | Yes | Yes |
| 2 | Resnick, H. H., et al. (2022). "Partial Recovery of Amblyopia After Fellow Eye Ischemic Optic Neuropathy." Journal of Neuro-Ophthalmology: 10.1097. | 0.88 | 0.88 | 0.00 | Unclear | 1.48 | 75 | 3 | Yes | Unclear |
| 3 | Resnick, H. H., et al. (2022). "Partial Recovery of Amblyopia After Fellow Eye Ischemic Optic Neuropathy." Journal of Neuro-Ophthalmology: 10.1097. | 0.30 | 0.10 | 2.00 | Unclear | 0.40 | 67 | 3 | Yes | Yes |
| 4 | Resnick, H. H., et al. (2022). "Partial Recovery of Amblyopia After Fellow Eye Ischemic Optic Neuropathy." Journal of Neuro-Ophthalmology: 10.1097. | 0.40 | 0.30 | 1.00 | 4 | 1.10 | 63 | 3 | No | Yes |
| 5 | Resnick, H. H., et al. (2022). "Partial Recovery of Amblyopia After Fellow Eye Ischemic Optic Neuropathy." Journal of Neuro-Ophthalmology: 10.1097. | 0.30 | 0.10 | 2.00 | 21 | 2.70 | 69 | 3 | Yes | Yes |
| 6 | Resnick, H. H., et al. (2022). "Partial Recovery of Amblyopia After Fellow Eye Ischemic Optic Neuropathy." Journal of Neuro-Ophthalmology: 10.1097. | 0.18 | 0.10 | 0.80 | 101 | 2.40 | 69 | 3 | Yes | Yes |
| 7 | Resnick, H. H., et al. (2022). "Partial Recovery of Amblyopia After Fellow Eye Ischemic Optic Neuropathy." Journal of Neuro-Ophthalmology: 10.1097. | 0.10 | 0.00 | 1.00 | 2 | 0.10 | 65 | 3 | No | Yes |
| 8 | Resnick, H. H., et al. (2022). "Partial Recovery of Amblyopia After Fellow Eye Ischemic Optic Neuropathy." Journal of Neuro-Ophthalmology: 10.1097. | 0.30 | 0.30 | 0.00 | Unclear | 2.10 | 69 | 3 | No | Unclear |
| 9 | Resnick, H. H., et al. (2022). "Partial Recovery of Amblyopia After Fellow Eye Ischemic Optic Neuropathy." Journal of Neuro-Ophthalmology: 10.1097. | 1.30 | 1.00 | 3.00 | 3 | 0.40 | 47 | 2 | No | Yes |
| 10 | Resnick, H. H., et al. (2022). "Partial Recovery of Amblyopia After Fellow Eye Ischemic Optic Neuropathy." Journal of Neuro-Ophthalmology: 10.1097. | 0.30 | 0.18 | 1.20 | 8 | 1.00 | 62 | 3 | Yes | Yes |
| 11 | Resnick, H. H., et al. (2022). "Partial Recovery of Amblyopia After Fellow Eye Ischemic Optic Neuropathy." Journal of Neuro-Ophthalmology: 10.1097. | 0.70 | 0.48 | 2.20 | 32 | 3.00 | 77 | 3 | Yes | Yes |
| 12 | Resnick, H. H., et al. (2022). "Partial Recovery of Amblyopia After Fellow Eye Ischemic Optic Neuropathy." Journal of Neuro-Ophthalmology: 10.1097. | 0.40 | 0.00 | 4.00 | 9 | 0.30 | 65 | 3 | No | Yes |
| 13 | Boase, A. J. Amblyopia, recovery at forty years of age. *Br J Ophthalmol* 35, 440–441 (1951). | 1.00 | 0.18 | 8.20 | 41 | 1.00 | 40 | 1 | Unclear | Yes |
| 14 | Constantinescu, C. S. & Gottlob, I. Possible role of corticosteroids in nervous system plasticity: improvement in amblyopia after optic neuritis in the fellow eye treated with steroids. Neurorehabil Neural Repair 15, 223–227 (2001). | 0.70 | 0.00 | 7.00 | 11 | 0.40 | 20 | 1 | Yes | Yes |
| 15 | Constantinescu, C. S. & Gottlob, I. Possible role of corticosteroids in nervous system plasticity: improvement in amblyopia after optic neuritis in the fellow eye treated with steroids. Neurorehabil Neural Repair 15, 223–227 (2001). | 0.16 | 0.00 | 1.60 | 1 | 0.70 | 22 | 1 | Yes | Yes |
| 16 | Constantinescu, C. S. & Gottlob, I. Possible role of corticosteroids in nervous system plasticity: improvement in amblyopia after optic neuritis in the fellow eye treated with steroids. Neurorehabil Neural Repair 15, 223–227 (2001). | 0.22 | 0.00 | 2.20 | 1 | 0.70 | 33 | 1 | No | Yes |
| 17 | Cynthia Douthwaite. Report on pleoptics. *British Orthoptic Journal* 17, 21–31 (1960). | 2.10 | 0.18 | 19.20 | 8 | 2.10 | 34 | 1 | Yes | No |
| 18 | Douthwaite, C. & Lee, B. An investigation of pleoptics. *British Orthoptic Journal* 15, 27–35 (1958). | 2.10 | 1.18 | 9.20 | Unclear | 1.00 | 46 | 2 | Yes | Yes |
| 19 | El Mallah, M. K. Amblyopia: is visual loss permanent? British Journal of Ophthalmology 84, 952–956 (2000). | 0.90 | 0.90 | 0.00 | Unclear | 0.70 | >61 | 3 | Yes | Unclear |
| 20 | El Mallah, M. K. Amblyopia: is visual loss permanent? British Journal of Ophthalmology 84, 952–956 (2000). | 1.50 | 1.20 | 3.00 | 12 | 0.80 | >61 | 3 | No | No |
| 21 | El Mallah, M. K. Amblyopia: is visual loss permanent? British Journal of Ophthalmology 84, 952–956 (2000). | 1.20 | 1.00 | 2.00 | 18 | 1.00 | >61 | 3 | No | No |
| 22 | El Mallah, M. K. Amblyopia: is visual loss permanent? British Journal of Ophthalmology 84, 952–956 (2000). | 1.20 | 0.80 | 4.00 | 6 | 1.20 | >61 | 3 | No | No |
| 23 | El Mallah, M. K. Amblyopia: is visual loss permanent? British Journal of Ophthalmology 84, 952–956 (2000). | 1.60 | 0.60 | 10.00 | 12 | 1.30 | >61 | 3 | No | No |
| 24 | El Mallah, M. K. Amblyopia: is visual loss permanent? British Journal of Ophthalmology 84, 952–956 (2000). | 1.60 | 1.00 | 6.00 | 12 | 1.30 | >61 | 3 | No | No |
| 25 | El Mallah, M. K. Amblyopia: is visual loss permanent? British Journal of Ophthalmology 84, 952–956 (2000). | 0.90 | 0.60 | 3.00 | 12 | 1.60 | >61 | 3 | No | No |
| 26 | El Mallah, M. K. Amblyopia: is visual loss permanent? British Journal of Ophthalmology 84, 952–956 (2000). | 1.60 | 1.30 | 3.00 | 24 | 1.60 | >61 | 3 | No | No |
| 27 | El Mallah, M. K. Amblyopia: is visual loss permanent? British Journal of Ophthalmology 84, 952–956 (2000). | 0.60 | 0.60 | 0.00 | Unclear | 1.60 | >61 | 3 | Yes | Unclear |
| 28 | Hamed et al; Improvement of vision in the contralateral Normal eye: a report of three cases. Binoc Vis (1991) | 0.60 | 0.30 | 3.00 | 1 | 0.54 | 70 | 3 | Yes | Yes |
| 29 | Hamed et al; Improvement of vision in the contralateral Normal eye: a report of three cases. Binoc Vis (1991) | 1.30 | 0.40 | 9.00 | 17 | 1.00 | 72 | 3 | Yes | Yes |
| 30 | Hamed et al; Improvement of vision in the contralateral Normal eye: a report of three cases. Binoc Vis (1991) | 1.00 | 0.00 | 10.00 | 5 | 2.40 | 54 | 2 | No | Yes |
| 31 | Hong, Y., Tao, Y., Shaojun, C. & Xiaohong, M. Visual improvement in an adult amblyopic eye following branch retinal vein occlusion in Non-amblyopic eye. Journal of Medical Colleges of PLA 27, 120–124 (2012). | 0.40 | 0.00 | 4.00 | 48 | 0.52 | 51 | 2 | No | No |
| 32 | Kaarniranta, K. & Kontkanen, M. Visual recovery of the amblyopic eye in an adult patient after loss of the dominant eye: A cta O phthalmologica S candinavica 2003. *Acta Ophthalmologica Scandinavica* 81, 539–539 (2003). | 0.52 | 0.00 | 5.20 | 26 | 3.00 | 19 | 1 | No | Yes |
| 33 | Karatza et al; Visual improvement in an adult amblyopic eye following radiation-induced visual loss in the contralateral eye. | 0.60 | 0.00 | 6.00 | 142 | 2.10 | 42 | 2 | No | No |
| 34 | Klaeger-Manzanell C, Hoyt CS, Good WV. Two step recovery of vision in the amblyopic eye after visual loss and enucleation of the fixing eye. Br J Ophthalmol. 1994;78:506-7. | 1.00 | 0.18 | 8.20 | 19 | 3.00 | 44 | 2 | Yes | Yes |
| 35 | Midgley, K. J., Miller, A. M., Ullman, M. A., Wells, K. & Fleischer, E. Cataract surgery in a moNocular amblyopic patient. JCRS Online Case Reports 6, 10–11 (2018). | 1.30 | 0.18 | 11.20 | Unclear | 3.00 | 67 | 3 | Unclear | Unclear |
| 36 | Rabin, J. Visual improvement in amblyopia after visual loss in the dominant eye. *Am J Optom Physiol Opt* 61, 334–337 (1984). | 1.00 | 0.18 | 8.20 | 36 | 1.30 | 52 | 2 | No | No |
| 37 | Tierney, D. W. Vision recovery in amblyopia after contralateral subretinal hemorrhage. *J Am Optom Assoc* 60, 281–283 (1989). | 1.00 | 0.18 | 8.20 | 11 | 1.78 | 64 | 3 | Yes | No |
| 38 | Tommila, V. & Tarkkanen, A. Treatment of amblyopia after loss of vision in the healthy eye. Ophthalmic Paediatrics and Genetics 1, 177–182 (1982). | 1.00 | 0.00 | 10.00 | Unclear | Not Available | 21-40 | 1 | Unclear | Unclear |
| 39 | Tommila, V. & Tarkkanen, A. Treatment of amblyopia after loss of vision in the healthy eye. Ophthalmic Paediatrics and Genetics 1, 177–182 (1982). | 1.30 | 0.30 | 10.00 | Unclear | Not Available | 21-40 | 1 | Unclear | Unclear |
| 40 | Tommila, V. & Tarkkanen, A. Treatment of amblyopia after loss of vision in the healthy eye. Ophthalmic Paediatrics and Genetics 1, 177–182 (1982). | 2.00 | 1.30 | 7.00 | Unclear | Not Available | 21-40 | 1 | Unclear | Unclear |
| 41 | Tommila, V. & Tarkkanen, A. Treatment of amblyopia after loss of vision in the healthy eye. Ophthalmic Paediatrics and Genetics 1, 177–182 (1982). | 0.82 | 0.40 | 4.20 | Unclear | Not Available | 21-40 | 1 | Unclear | Unclear |
| 42 | Tommila, V. & Tarkkanen, A. Treatment of amblyopia after loss of vision in the healthy eye. Ophthalmic Paediatrics and Genetics 1, 177–182 (1982). | 1.00 | 0.52 | 4.80 | Unclear | Not Available | 21-40 | 1 | Unclear | Unclear |
| 43 | Tommila, V. & Tarkkanen, A. Treatment of amblyopia after loss of vision in the healthy eye. Ophthalmic Paediatrics and Genetics 1, 177–182 (1982). | 1.00 | 0.52 | 4.80 | Unclear | Not Available | 41-60 | 2 | Unclear | Unclear |
| 44 | Tommila, V. & Tarkkanen, A. Treatment of amblyopia after loss of vision in the healthy eye. Ophthalmic Paediatrics and Genetics 1, 177–182 (1982). | 1.00 | 0.40 | 6.00 | Unclear | Not Available | 41-60 | 2 | Unclear | Unclear |
| 45 | Tommila, V. & Tarkkanen, A. Treatment of amblyopia after loss of vision in the healthy eye. Ophthalmic Paediatrics and Genetics 1, 177–182 (1982). | 0.70 | 0.30 | 4.00 | Unclear | Not Available | 41-60 | 2 | Unclear | Unclear |
| 46 | Tommila, V. & Tarkkanen, A. Treatment of amblyopia after loss of vision in the healthy eye. Ophthalmic Paediatrics and Genetics 1, 177–182 (1982). | 0.40 | 0.00 | 4.00 | Unclear | Not Available | 41-60 | 2 | Unclear | Unclear |
| 47 | Tommila, V. & Tarkkanen, A. Treatment of amblyopia after loss of vision in the healthy eye. Ophthalmic Paediatrics and Genetics 1, 177–182 (1982). | 0.82 | 0.40 | 4.20 | Unclear | Not Available | >61 | 3 | Unclear | Unclear |
| 48 | Tommila, V. TREATMENT OF AMBLYOPIA IN LOSS OF VISION IN THE OTHER EYE. Acta Ophthalmol (Copenh) 42, 489–494 (1964). | 1.00 | 0.00 | 10.00 | 1 | 3.00 | 32 | 1 | No | Yes |
| 49 | Vagge, A., Shields, C. L., Shields, J. A., Pointdujour-Lim, R. & Schnall, B. Visual improvement in amblyopic eye following treatment-induced vision loss in dominant eye with uveal melaNoma. Br J Ophthalmol 104, 202–207 (2020). | 1.00 | 0.70 | 3.00 | 12 | 0.30 | 73 | 3 | Unclear | Unclear |
| 50 | Vagge, A., Shields, C. L., Shields, J. A., Pointdujour-Lim, R. & Schnall, B. Visual improvement in amblyopic eye following treatment-induced vision loss in dominant eye with uveal melaNoma. Br J Ophthalmol 104, 202–207 (2020). | 0.60 | 0.54 | 0.60 | 48 | 0.30 | 72 | 3 | Unclear | Unclear |
| 51 | Vagge, A., Shields, C. L., Shields, J. A., Pointdujour-Lim, R. & Schnall, B. Visual improvement in amblyopic eye following treatment-induced vision loss in dominant eye with uveal melaNoma. Br J Ophthalmol 104, 202–207 (2020). | 0.70 | 0.70 | 0.00 | Unclear | 0.30 | 68 | 3 | Unclear | Unclear |
| 52 | Vagge, A., Shields, C. L., Shields, J. A., Pointdujour-Lim, R. & Schnall, B. Visual improvement in amblyopic eye following treatment-induced vision loss in dominant eye with uveal melaNoma. Br J Ophthalmol 104, 202–207 (2020). | 0.40 | 0.10 | 3.00 | 24 | 0.30 | 55 | 2 | Unclear | Unclear |
| 53 | Vagge, A., Shields, C. L., Shields, J. A., Pointdujour-Lim, R. & Schnall, B. Visual improvement in amblyopic eye following treatment-induced vision loss in dominant eye with uveal melaNoma. Br J Ophthalmol 104, 202–207 (2020). | 0.40 | 0.30 | 1.00 | 5 | 0.54 | 49 | 2 | Unclear | Unclear |
| 54 | Vagge, A., Shields, C. L., Shields, J. A., Pointdujour-Lim, R. & Schnall, B. Visual improvement in amblyopic eye following treatment-induced vision loss in dominant eye with uveal melaNoma. Br J Ophthalmol 104, 202–207 (2020). | 0.60 | 0.00 | 6.00 | 12 | 0.60 | 50 | 2 | Unclear | Unclear |
| 55 | Vagge, A., Shields, C. L., Shields, J. A., Pointdujour-Lim, R. & Schnall, B. Visual improvement in amblyopic eye following treatment-induced vision loss in dominant eye with uveal melaNoma. Br J Ophthalmol 104, 202–207 (2020). | 1.30 | 1.00 | 3.00 | 12 | 0.60 | 71 | 3 | Unclear | Unclear |
| 56 | Vagge, A., Shields, C. L., Shields, J. A., Pointdujour-Lim, R. & Schnall, B. Visual improvement in amblyopic eye following treatment-induced vision loss in dominant eye with uveal melaNoma. Br J Ophthalmol 104, 202–207 (2020). | 2.10 | 2.10 | 0.00 | Unclear | 0.70 | 55 | 2 | Unclear | Unclear |
| 57 | Vagge, A., Shields, C. L., Shields, J. A., Pointdujour-Lim, R. & Schnall, B. Visual improvement in amblyopic eye following treatment-induced vision loss in dominant eye with uveal melaNoma. Br J Ophthalmol 104, 202–207 (2020). | 0.48 | 0.10 | 3.80 | 36 | 1.00 | 49 | 2 | Unclear | Unclear |
| 58 | Vagge, A., Shields, C. L., Shields, J. A., Pointdujour-Lim, R. & Schnall, B. Visual improvement in amblyopic eye following treatment-induced vision loss in dominant eye with uveal melaNoma. Br J Ophthalmol 104, 202–207 (2020). | 1.30 | 0.70 | 6.00 | 108 | 1.18 | 63 | 3 | Unclear | Unclear |
| 59 | Vagge, A., Shields, C. L., Shields, J. A., Pointdujour-Lim, R. & Schnall, B. Visual improvement in amblyopic eye following treatment-induced vision loss in dominant eye with uveal melaNoma. Br J Ophthalmol 104, 202–207 (2020). | 0.30 | 0.00 | 3.00 | 24 | 2.10 | 40 | 1 | Unclear | Unclear |
| 60 | Vagge, A., Shields, C. L., Shields, J. A., Pointdujour-Lim, R. & Schnall, B. Visual improvement in amblyopic eye following treatment-induced vision loss in dominant eye with uveal melaNoma. Br J Ophthalmol 104, 202–207 (2020). | 1.00 | 1.00 | 0.00 | Unclear | 2.10 | 39 | 1 | Unclear | Unclear |
| 61 | Vagge, A., Shields, C. L., Shields, J. A., Pointdujour-Lim, R. & Schnall, B. Visual improvement in amblyopic eye following treatment-induced vision loss in dominant eye with uveal melaNoma. Br J Ophthalmol 104, 202–207 (2020). | 0.30 | 0.18 | 1.20 | 12 | 2.10 | 43 | 2 | Unclear | Unclear |
| 62 | Vagge, A., Shields, C. L., Shields, J. A., Pointdujour-Lim, R. & Schnall, B. Visual improvement in amblyopic eye following treatment-induced vision loss in dominant eye with uveal melaNoma. Br J Ophthalmol 104, 202–207 (2020). | 0.54 | 0.00 | 5.40 | 12 | 2.10 | 66 | 3 | Unclear | Unclear |
| 63 | Vagge, A., Shields, C. L., Shields, J. A., Pointdujour-Lim, R. & Schnall, B. Visual improvement in amblyopic eye following treatment-induced vision loss in dominant eye with uveal melaNoma. Br J Ophthalmol 104, 202–207 (2020). | 0.60 | 0.10 | 5.00 | 144 | 2.10 | 70 | 3 | Unclear | Unclear |
| 64 | Vagge, A., Shields, C. L., Shields, J. A., Pointdujour-Lim, R. & Schnall, B. Visual improvement in amblyopic eye following treatment-induced vision loss in dominant eye with uveal melaNoma. Br J Ophthalmol 104, 202–207 (2020). | 0.60 | 0.88 | -2.80 | Unclear | 2.10 | 56 | 2 | Unclear | Unclear |
| 65 | Vagge, A., Shields, C. L., Shields, J. A., Pointdujour-Lim, R. & Schnall, B. Visual improvement in amblyopic eye following treatment-induced vision loss in dominant eye with uveal melaNoma. Br J Ophthalmol 104, 202–207 (2020). | 0.60 | 0.00 | 6.00 | 108 | 2.10 | 42 | 2 | Unclear | Unclear |
| 66 | Vagge, A., Shields, C. L., Shields, J. A., Pointdujour-Lim, R. & Schnall, B. Visual improvement in amblyopic eye following treatment-induced vision loss in dominant eye with uveal melaNoma. Br J Ophthalmol 104, 202–207 (2020). | 2.10 | 1.18 | 9.20 | 12 | 2.40 | 62 | 3 | Unclear | Unclear |
| 67 | Vagge, A., Shields, C. L., Shields, J. A., Pointdujour-Lim, R. & Schnall, B. Visual improvement in amblyopic eye following treatment-induced vision loss in dominant eye with uveal melaNoma. Br J Ophthalmol 104, 202–207 (2020). | 0.30 | 0.18 | 1.20 | 12 | 2.40 | 57 | 2 | Unclear | Unclear |
| 68 | Vagge, A., Shields, C. L., Shields, J. A., Pointdujour-Lim, R. & Schnall, B. Visual improvement in amblyopic eye following treatment-induced vision loss in dominant eye with uveal melaNoma. Br J Ophthalmol 104, 202–207 (2020). | 0.30 | 0.00 | 3.00 | 12 | 2.70 | 52 | 2 | Unclear | Unclear |
| 69 | Vagge, A., Shields, C. L., Shields, J. A., Pointdujour-Lim, R. & Schnall, B. Visual improvement in amblyopic eye following treatment-induced vision loss in dominant eye with uveal melaNoma. Br J Ophthalmol 104, 202–207 (2020). | 0.48 | 0.00 | 4.80 | 12 | 3.00 | 54 | 2 | Unclear | Unclear |
| 70 | van de Graaf, E. S., Despriet, D. D. G., Klaver, C. C. W. & Simonsz, H. J. Patient-reported utilities in bilateral visual impairment from amblyopia and age-related macular degeneration. *BMC Ophthalmol* 16, 56 (2016). | 0.10 | 0.04 | 0.60 | Unclear | Not Available | Unclear | Unclear | Unclear | No |
| 71 | Vereecken, E. P. & Brabant, P. ProgNosis for Vision in Amblyopia After the Loss of the Good Eye. Archives of Ophthalmology 102, 220–224 (1984). | 0.40 | 0.04 | 3.60 | 1 | Not Available | 24 | 1 | Unclear | Unclear |
| 72 | Vereecken, E. P. & Brabant, P. ProgNosis for Vision in Amblyopia After the Loss of the Good Eye. Archives of Ophthalmology 102, 220–224 (1984). | 0.70 | 0.04 | 6.60 | 12 | Not Available | 32 | 1 | Unclear | Unclear |
| 73 | Vereecken, E. P. & Brabant, P. ProgNosis for Vision in Amblyopia After the Loss of the Good Eye. Archives of Ophthalmology 102, 220–224 (1984). | 0.40 | 0.04 | 3.60 | 12 | Not Available | 51 | 2 | Unclear | Unclear |
| 74 | Vereecken, E. P. & Brabant, P. ProgNosis for Vision in Amblyopia After the Loss of the Good Eye. Archives of Ophthalmology 102, 220–224 (1984). | 0.52 | 0.00 | 5.20 | 12 | Not Available | 23 | 1 | Unclear | Unclear |
| 75 | Vereecken, E. P. & Brabant, P. ProgNosis for Vision in Amblyopia After the Loss of the Good Eye. Archives of Ophthalmology 102, 220–224 (1984). | 0.40 | 0.00 | 4.00 | 12 | Not Available | 41 | 2 | Unclear | Unclear |
| 76 | Vereecken, E. P. & Brabant, P. ProgNosis for Vision in Amblyopia After the Loss of the Good Eye. Archives of Ophthalmology 102, 220–224 (1984). | 1.00 | 0.10 | 9.00 | 12 | Not Available | 57 | 2 | Unclear | Unclear |
| 77 | Vereecken, E. P. & Brabant, P. ProgNosis for Vision in Amblyopia After the Loss of the Good Eye. Archives of Ophthalmology 102, 220–224 (1984). | 1.00 | 0.10 | 9.00 | 24 | Not Available | 22 | 1 | Unclear | Unclear |
| 78 | Vereecken, E. P. & Brabant, P. ProgNosis for Vision in Amblyopia After the Loss of the Good Eye. Archives of Ophthalmology 102, 220–224 (1984). | 1.00 | 0.10 | 9.00 | 120 | Not Available | 23 | 1 | Unclear | Unclear |
| 79 | Vereecken, E. P. & Brabant, P. ProgNosis for Vision in Amblyopia After the Loss of the Good Eye. Archives of Ophthalmology 102, 220–224 (1984). | 1.00 | 0.22 | 7.80 | 2 | Not Available | 77 | 3 | Unclear | Unclear |
| 80 | Vereecken, E. P. & Brabant, P. ProgNosis for Vision in Amblyopia After the Loss of the Good Eye. Archives of Ophthalmology 102, 220–224 (1984). | 0.70 | 0.22 | 4.80 | 6 | Not Available | 50 | 2 | Unclear | Unclear |
| 81 | Vereecken, E. P. & Brabant, P. ProgNosis for Vision in Amblyopia After the Loss of the Good Eye. Archives of Ophthalmology 102, 220–224 (1984). | 1.00 | 0.30 | 7.00 | 24 | Not Available | 22 | 1 | Unclear | Unclear |
| 82 | Vereecken, E. P. & Brabant, P. ProgNosis for Vision in Amblyopia After the Loss of the Good Eye. Archives of Ophthalmology 102, 220–224 (1984). | 0.52 | 0.22 | 3.00 | 12 | Not Available | 52 | 2 | Unclear | Unclear |
| 83 | Vereecken, E. P. & Brabant, P. ProgNosis for Vision in Amblyopia After the Loss of the Good Eye. Archives of Ophthalmology 102, 220–224 (1984). | 0.52 | 0.22 | 3.00 | 40 | Not Available | 35 | 1 | Unclear | Unclear |
| 84 | Vereecken, E. P. & Brabant, P. ProgNosis for Vision in Amblyopia After the Loss of the Good Eye. Archives of Ophthalmology 102, 220–224 (1984). | 1.00 | 0.22 | 7.80 | 4 | Not Available | 40 | 1 | Unclear | Unclear |
| 85 | Vereecken, E. P. & Brabant, P. ProgNosis for Vision in Amblyopia After the Loss of the Good Eye. Archives of Ophthalmology 102, 220–224 (1984). | 1.00 | 0.22 | 7.80 | 7 | Not Available | 58 | 2 | Unclear | Unclear |
| 86 | Vereecken, E. P. & Brabant, P. ProgNosis for Vision in Amblyopia After the Loss of the Good Eye. Archives of Ophthalmology 102, 220–224 (1984). | 1.00 | 0.22 | 7.80 | 10 | Not Available | 37 | 1 | Unclear | Unclear |
| 87 | Vereecken, E. P. & Brabant, P. ProgNosis for Vision in Amblyopia After the Loss of the Good Eye. Archives of Ophthalmology 102, 220–224 (1984). | 1.00 | 0.22 | 7.80 | 36 | Not Available | 26 | 1 | Unclear | Unclear |
| 88 | Vereecken, E. P. & Brabant, P. ProgNosis for Vision in Amblyopia After the Loss of the Good Eye. Archives of Ophthalmology 102, 220–224 (1984). | 0.70 | 0.16 | 5.40 | 6 | Not Available | 19 | 1 | Unclear | Unclear |
| 89 | Vereecken, E. P. & Brabant, P. ProgNosis for Vision in Amblyopia After the Loss of the Good Eye. Archives of Ophthalmology 102, 220–224 (1984). | 1.00 | 0.16 | 8.40 | 72 | Not Available | 18 | 1 | Unclear | Unclear |
| 90 | Vereecken, E. P. & Brabant, P. ProgNosis for Vision in Amblyopia After the Loss of the Good Eye. Archives of Ophthalmology 102, 220–224 (1984). | 1.00 | 0.22 | 7.80 | 6 | Not Available | 68 | 3 | Unclear | Unclear |
| 91 | Vereecken, E. P. & Brabant, P. ProgNosis for Vision in Amblyopia After the Loss of the Good Eye. Archives of Ophthalmology 102, 220–224 (1984). | 1.00 | 0.16 | 8.40 | Unclear | Not Available | 37 | 1 | Unclear | Unclear |
| 92 | Vereecken, E. P. & Brabant, P. ProgNosis for Vision in Amblyopia After the Loss of the Good Eye. Archives of Ophthalmology 102, 220–224 (1984). | 1.00 | 0.22 | 7.80 | 18 | Not Available | 52 | 2 | Unclear | Unclear |
| 93 | Vereecken, E. P. & Brabant, P. ProgNosis for Vision in Amblyopia After the Loss of the Good Eye. Archives of Ophthalmology 102, 220–224 (1984). | 1.00 | 0.22 | 7.80 | 12 | Not Available | 49 | 2 | Unclear | Unclear |
| 94 | Vereecken, E. P. & Brabant, P. ProgNosis for Vision in Amblyopia After the Loss of the Good Eye. Archives of Ophthalmology 102, 220–224 (1984). | 1.00 | 0.22 | 7.80 | Unclear | Not Available | 60 | 2 | Unclear | Unclear |
| 95 | Vereecken, E. P. & Brabant, P. ProgNosis for Vision in Amblyopia After the Loss of the Good Eye. Archives of Ophthalmology 102, 220–224 (1984). | 1.00 | 0.30 | 7.00 | 12 | Not Available | 34 | 1 | Unclear | Unclear |
| 96 | Vereecken, E. P. & Brabant, P. ProgNosis for Vision in Amblyopia After the Loss of the Good Eye. Archives of Ophthalmology 102, 220–224 (1984). | 1.00 | 0.16 | 8.40 | 6 | Not Available | 65 | 3 | Unclear | Unclear |
| 97 | Vereecken, E. P. & Brabant, P. ProgNosis for Vision in Amblyopia After the Loss of the Good Eye. Archives of Ophthalmology 102, 220–224 (1984). | 1.00 | 0.30 | 7.00 | 13 | Not Available | 63 | 3 | Unclear | Unclear |
| 98 | Vereecken, E. P. & Brabant, P. ProgNosis for Vision in Amblyopia After the Loss of the Good Eye. Archives of Ophthalmology 102, 220–224 (1984). | 1.00 | 0.30 | 7.00 | 6 | Not Available | 34 | 1 | Unclear | Unclear |
| 99 | Vereecken, E. P. & Brabant, P. ProgNosis for Vision in Amblyopia After the Loss of the Good Eye. Archives of Ophthalmology 102, 220–224 (1984). | 1.00 | 0.40 | 6.00 | 18 | Not Available | 54 | 2 | Unclear | Unclear |
| 100 | Vereecken, E. P. & Brabant, P. ProgNosis for Vision in Amblyopia After the Loss of the Good Eye. Archives of Ophthalmology 102, 220–224 (1984). | 1.00 | 0.44 | 5.60 | 6 | Not Available | 39 | 1 | Unclear | Unclear |
| 101 | Vereecken, E. P. & Brabant, P. ProgNosis for Vision in Amblyopia After the Loss of the Good Eye. Archives of Ophthalmology 102, 220–224 (1984). | 1.00 | 0.52 | 4.80 | Unclear | Not Available | 60 | 2 | Unclear | Unclear |
| 102 | Vereecken, E. P. & Brabant, P. ProgNosis for Vision in Amblyopia After the Loss of the Good Eye. Archives of Ophthalmology 102, 220–224 (1984). | 1.00 | 0.54 | 4.60 | 12 | Not Available | 20 | 1 | Unclear | Unclear |
| 103 | Vereecken, E. P. & Brabant, P. ProgNosis for Vision in Amblyopia After the Loss of the Good Eye. Archives of Ophthalmology 102, 220–224 (1984). | 1.00 | 0.52 | 4.80 | Unclear | Not Available | 30 | 1 | Unclear | Unclear |
| 104 | Vereecken, E. P. & Brabant, P. ProgNosis for Vision in Amblyopia After the Loss of the Good Eye. Archives of Ophthalmology 102, 220–224 (1984). | 1.00 | 0.40 | 6.00 | Unclear | Not Available | 60 | 2 | Unclear | Unclear |
| 105 | Wilson ME. Adult amblyopia reversed by contralateral cataract formation. J Pediatr Ophthalmol Strabismus. 1992;29:100-2. | 0.70 | 0.30 | 4.00 | 48 | 1.30 | 63 | 3 | Yes | No |
| 106 | Wilson ME. Adult amblyopia reversed by contralateral cataract formation. J Pediatr Ophthalmol Strabismus. 1992;29:100-2. | 0.48 | 0.10 | 3.80 | 12 | 2.40 | 51 | 2 | Yes | No |
| 107 | Stringer, E. A case of eccentric fixation in an adult in which central fixation hase been restored. *British Orthoptic Journal* 14, 61–68 (1957). | 2.10 | 0.60 | 15.00 | 12 | 3.00 | 51 | 2 | Yes | Yes |
| 108 | Douthwaite, C. M. Orthoptics in Lyons. *British Orthoptic Journal* 14, 56–60 (1957). | 1.00 | 0.60 | 4.00 | Unclear | Not Available | 54 | 2 | Yes | Unclear |
| 109 | Orge Faruk H. & Dar Suhail A. Visual Acuity Improvement of Amblyopia in an Adult With Levodopa/Carbidopa Treatment. *Journal of Pediatric Ophthalmology & Strabismus* 52, e45–e47 (2015). | 0.40 | 0.18 | 2.20 | Unclear | 3.00 | 41 | 2 | No | Yes |
